# Brain-wide pleiotropy investigation of alcohol drinking and tobacco smoking behaviors

**DOI:** 10.1101/2024.05.27.24307989

**Authors:** Giovanni Deiana, Jun He, Brenda Cabrera-Mendoza, Roberto Ciccocioppo, Valerio Napolioni, Renato Polimanti

**Author notes:** **Correspondence:** Renato Polimanti, PhD. Department of Psychiatry, Yale University School of Medicine. 60 Temple, Suite 7A, New Haven, CT 06511, USA.

## Abstract

To investigate the pleiotropic mechanisms linking brain structure and function to alcohol drinking and tobacco smoking, we integrated genome-wide data generated by the GWAS and Sequencing Consortium of Alcohol and Nicotine use (GSCAN; up to 805,431 participants) with information related to 3,935 brain imaging-derived phenotypes (IDPs) available from UK Biobank (N=33,224). We observed global genetic correlation of smoking behaviors with white matter hyperintensities, the morphology of the superior longitudinal fasciculus, and the mean thickness of pole-occipital. With respect to the latter brain IDP, we identified a local genetic correlation with age at which the individual began smoking regularly (hg38 chr2:35,895,678-36,640,246: rho=1, p=1.01×10^−5^). This region has been previously associated with smoking initiation, educational attainment, chronotype, and cortical thickness. Our genetically informed causal inference analysis using both latent causal variable approach and Mendelian randomization linked the activity of prefrontal and premotor cortex and that of superior and inferior precentral sulci, and cingulate sulci to the number of alcoholic drinks per week (genetic causality proportion, gcp=0.38, p=8.9×10^−4^, rho=-0.18±0.07; inverse variance weighting, IVW beta=-0.04, 95%CI=-0.07 – −0.01). This relationship could be related to the role of these brain regions in the modulation of reward-seeking motivation and the processing of social cues. Overall, our brain-wide investigation highlighted that different pleiotropic mechanisms likely contribute to the relationship of brain structure and function with alcohol drinking and tobacco smoking, suggesting decision-making activities and chemosensory processing as modulators of propensity towards alcohol and tobacco consumption.

## INTRODUCTION

Alcohol drinking and tobacco smoking are among the leading causes of death worldwide due to their high prevalence [1,2]. This widespread use is partially attributed to the complex interplay of alcohol and nicotine with human brain, as shown by previous studies [3–5]. Additionally, several analyses observed consistent pleiotropy linking alcohol drinking and tobacco smoking to brain structure and function. For instance, early tobacco smoking initiation was genetically correlated with an increased precuneus surface area and decreased cortical thickness and surface area of the inferior temporal gyrus [6]. Similarly, alcohol drinking showed genetic correlation with an increased total cortical surface area and decreased average cortical thickness [6]. Because genetic information can be used as an anchor for causal inference [7], investigators also explored possible direct effects between drinking/smoking behaviors and brain morphology. For example, smoking initiation and alcohol drinking appear to have a possible causal association with decreased gray matter volume and the multivariable analysis pointed to alcohol drinking as the potential primary driver of this relationship [8]. In another study, genetically predicted global cortical thickness showed an effect on alcohol drinking behaviors that was independent of neuropsychiatric phenotypes, substance use, trauma, and neurodegeneration [9]. Focusing on specific hypotheses, these previous investigations advanced our understanding of the brain mechanisms contributing to alcohol drinking and tobacco smoking behaviors. However, large-scale datasets allow investigators to expand further the depth of the analyses. Indeed, recent brain-wide pleiotropy analyses provided new insights into the role of brain structure and function on neuropsychiatric and behavioral traits [10–13].

In the present study, we systematically investigated the pleiotropic mechanisms linking alcohol drinking and tobacco smoking to brain structure and function. Specifically, we integrated genome-wide data generated by the genome-wide association studies (GWAS) and Sequencing Consortium of Alcohol and Nicotine use (GSCAN; up to 805,431 participants) [14] with information related to 3,935 brain imaging-derived phenotypes (IDPs; Supplemental Table 1) obtained from six magnetic resonance imaging (MRI) modalities [15], exploring the contribution of different pleiotropic mechanisms in the interplay between drinking/smoking behaviors and human brain.

## MATERIALS AND METHODS

### Study Design

In the present study, we investigated the pleiotropic mechanisms linking alcohol drinking and tobacco smoking behaviors to brain structure and function by applying multiple analytic approaches to large-scale genome-wide datasets (Figure 1). A global genetic correlation analysis was performed to assess the overall genetic overlap between alcohol drinking and tobacco smoking and brain IDPs. Considering global genetic correlations surviving multiple testing correction, we leveraged the local analysis of [co]variant association (LAVA) approach [16] to identify chromosomal regions with strong statistical evidence of shared genetic mechanisms. To assess the presence of causal relationships underlying the global genetic overlap observed, we applied the latent causal variable (LCV) approach [17] to pairwise combinations of brain IDP and alcohol/tobacco-related behaviors reaching nominally significant genetic correlation. Mendelian Randomization (MR) analysis was also conducted to follow up on the false discovery rate (FDR) significant results obtained from the LCV analysis.

**Fig. 1:**
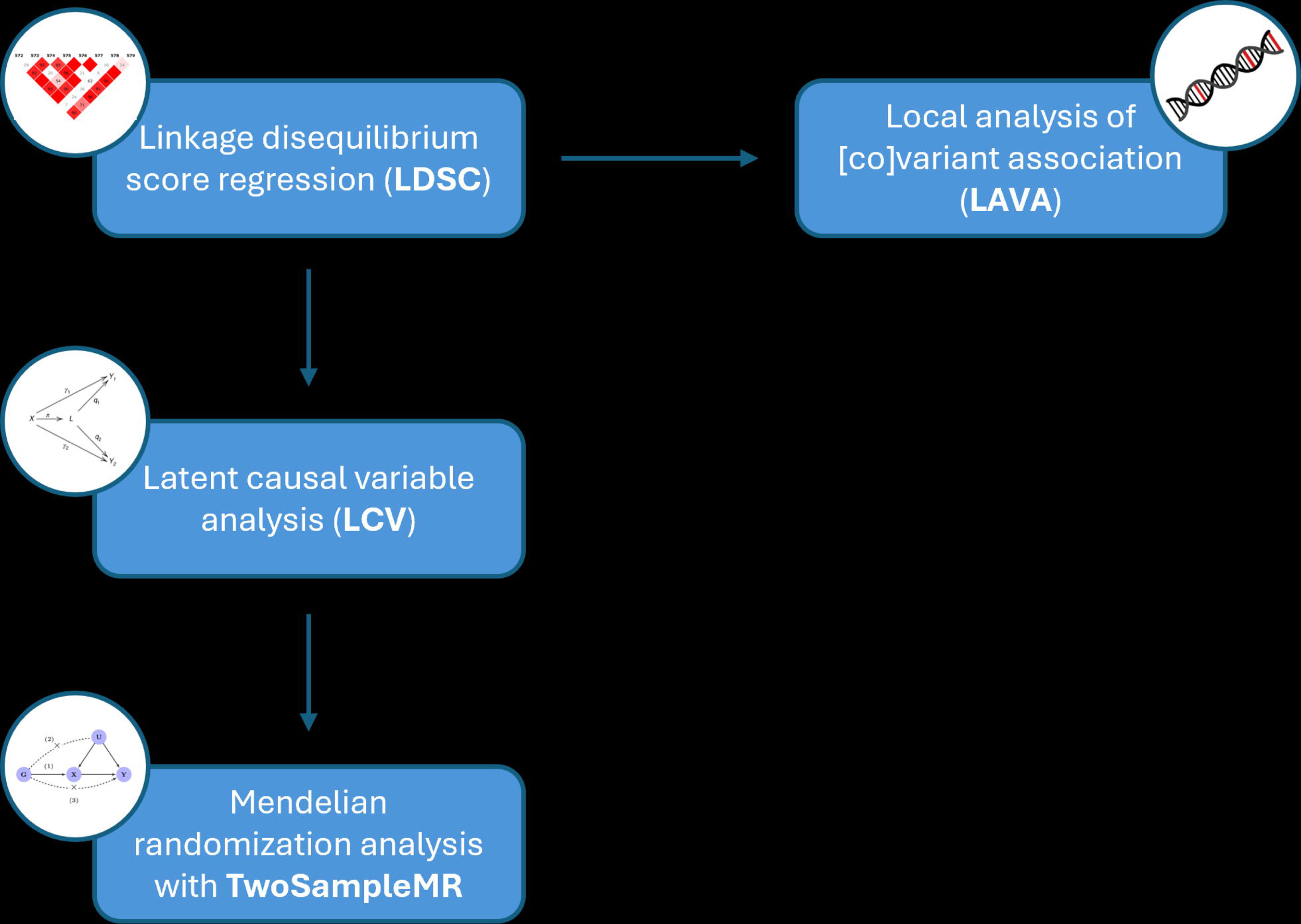
Workflow of the analyses performed.

### Data sources

Genome-wide association statistics regarding IDPs were derived from the UK Biobank (UKB). The UKB is a large population-based prospective cohort containing in-depth genetic and health information from over 500,000 participants [18]. In UKB, brain imaging was conducted using six MRI modalities: T1-weighted structural image, T2-weighted fluid-attenuated inversion recovery (T2_FLAIR) structural image, diffusion MRI (dMRI), resting-state functional MRI (rfMRI), task functional MRI (tfMRI), and susceptibility-weighted imaging (SWI). A total of 3,935 brain IDPs (Supplemental Table 1) were defined from MRI scans [19]. GWAS of brain IDPs in up to 33,224 UKB participants of European descent was previously described [15].

Genome-wide association statistics regarding behaviors related to alcohol drinking and tobacco smoking were derived from GSCAN. In its latest GWAS [14], this large collaborative effort meta-analyzed genome-wide information regarding smoking initiation (SmkInit) and the age at which the individual began smoking regularly (AgeSmk), cigarettes smoked per day (CigDay), smoking cessation (SmkCes), and alcoholic drinks per week (DrnkWk). Since brain IDP GWAS data were available only for individuals of European descent, we used publicly available GSCAN GWAS data for the same ancestry group (SmkInit N=805,431; AgeSmk N=323,386; CigDay N=326,497; SmkCes N= 388,313; DrnkWk N= 666,978). The sample overlap due to UKB inclusion in both IDP and GSCAN GWAS does not affect genetic correlation, LAVA, and LCV analyses. However, to avoid potential sample overlap bias in Mendelian randomization (MR) analysis, we also analyzed GSCAN GWAS data excluding the UKB cohort (SmkInit N= 357,235; AgeSmk N= 175,835; CigDay N= 183,196; SmkCes N= 188,701; DrnkWk N= 304,322).

### Linkage disequilibrium score regression

Single nucleotide polymorphism (SNP)-based heritability and global genetic correlation were estimated using the linkage disequilibrium score regression (LDSC) method [20]. These analyses were performed using the HapMap 3 reference panel [21] and LD scores derived from European reference populations available from the 1000 Genomes Project [22]. Statistically significant genetic correlations were determined considering FDR q<0.05 to account for the number of brain IDPs tested with respect to each GSCAN phenotype.

### Local analysis of [co]variant association

Local genetic correlation was assessed across 2,495 semi-independent chromosomal regions (∼1Mb window) using LAVA [16]. The LAVA univariate analysis was performed to estimate local SNP-based heritability for each pair of GSCAN phenotype and brain IDP. Considering chromosomal regions with at least nominally significant local SNP-based heritability (p<0.05), we estimated local genetic correlation between GSCAN phenotypes and brain IDPs using LAVA bivariate analysis. FDR multiple testing correction (FDR q<0.05) accounting for the number of chromosomal regions tested was applied to define statistically significant local genetic correlations. To further characterize the genomic regions identified using the LAVA approach, we leveraged information available from the GWAS catalog including genetic regions and reports from previous associations [23].

### Genetically Inferred Causal Inference

We performed a LCV analysis [17] to estimate whether the global genetic correlation observed between GSCAN phenotypes and brain IDPs was due to possible cause-effect relationships. Considering pair combinations that reached at least nominally significance in the LDSC genetic correlation analysis (p<0.05), we estimated the genetic causality proportions (gcp) between two traits. The gcp statistics can range from −1 to 1, where gcp=0 indicates no genetic causality, gcp=1 indicates a full genetic causality of trait #1 on trait #2, and gcp=-1 indicates full genetic causality of trait #2 on trait #1. In the present study, LCV analyses were performed considering brain IDPs phenotypes as trait #1 and GSCAN phenotypes as trait #2. Accordingly, positive gcp estimates indicate a causal effect of brain IDPs on GSCAN phenotypes, while a negative gcp estimate indicates a causal effect in the reverse direction. The sign of the genetically inferred causal effect is defined by the sign of the LCV rho statistics (i.e., rho>0 corresponds to positive causal effects, while rho<0 corresponds to negative causal effects). FDR correction accounting for the number of tests performed (FDR q<0.05) was applied to define statistically significant LCV results.

To complement LCV results, we performed a MR analysis. Although both methods evaluate causal effect relationships, the LCV and MR results are based on different assumptions. Accordingly, an effect consistent between these two approaches can be considered more reliable. The MR analysis was performed using the TwoSampleMR package [24] to estimate inverse variance weighting (IVW) estimates. Independent genetic instruments for TwoSampleMR analyses were identified considering SNPs with an exposure GWAS P-value threshold of 1×10^−5^, that were LD-independent (r^2^=0.001 within a 10,000-kb window). Because MR analyses can be biased by sample overlap, this analysis was conducted using UKB brain-IDP GWAS and GSCAN GWAS data excluding UKB participants.

## RESULTS

After FDR multiple testing correction (FDR q<0.05), we identified three genetic correlations linking behaviors related to tobacco smoking with brain structure and function (Supplemental Table 2). AgeSmk showed a negative genetic correlation with the mean thickness of Pole-occipital in the left hemisphere generated by Destrieux (a2009s) parcellation of the white surface (aparc-a2009s lh thickness Pole-occipital, IDP 1219; rg=-0.23, p=7.74×10^−6^). A positive genetic correlation was observed between SmCes and the total volume of white matter hyperintensities from T1 and T2_FLAIR images (T2 FLAIR BIANCA WMH volume, IDP 1437; rg=0.16, p=1.03×10^−5^). CigDay also showed a positive correlation with the mean second level (L2) of right superior longitudinal fasciculus on fractional anisotropy skeleton from dMRI data (dMRI TBSS L2 Superior longitudinal fasciculus R, IDP 1736; rg=0.14, p=1.24×10^−5^). To further investigate the dynamics underlying these relationships, we conducted a local genetic correlation analysis and identified one region (hg38 chr2:35,895,678-36,640,246) showing statistically significant local genetic correlation between AgeSmk and aparc-a2009s lh thickness Pole-occipital (rho=1, p=1.01×10^−5^). In this region, 178 genome-wide significant associations (p<5X10^−8^) were reported in the GWAS catalog [23] (Supplemental Table 3). Among them, several were related to brain related phenotypes including smoking initiation (rs62134085 p=1×10^−18^), educational attainment (rs305191 p=2×10^−14^), chronotype (rs848552 p=5×10^−14^), self-reported math ability (rs6708545 p=1×10^−10^), cognitive performance (rs6728742 p=1×10^−8^), and cortical thickness (rs1017154 p=3×10^−8^).

As mentioned above, positive gcp estimates in the LCV analysis indicate a causal effect of brain IDPs on GSCAN phenotypes, while the sign of the genetically inferred causal effect is defined by the sign of the LCV rho statistics (i.e., rho>0 corresponds to positive causal effects, while rho<0 corresponds to negative causal effects). Considering nominally significant genetic correlations between brain IDPs and GSCAN phenotypes (Supplemental Table 2), we found 19 relationships linking brain structure and function to behaviors related to alcohol drinking and tobacco smoking (gcp>0, FDR q<0.05; Figure 2, Supplemental Table 4). Among them, 12 were related to brain connectivity analysis derived from rfMRI: three were related to DrnkWk (e.g, partial correlation of edge 363 in rfMRI dimensionality 100, ICA100 edge 363, IDP 2791; gcp=0.77, P=1.12×10^−16^, rho=0.18±0.07), four to AgeSmk (e.g., ICA100 edge 838, IDP 3266, Supplemental Figure 1; gcp=0.79, P=3.72×10^−23^, rho=0.20±0.08), three to CigDay (e.g., ICA25 edge 184, IDP 2402, Supplemental Figure 2; gcp=0.47, P=2.13×10^−20^, rho=-0.20±0.08), and two to SmkCes (e.g., ICA25 edge 190, IDP 2408; gcp=0.86, P=9.43×10^−22^, rho=0.21±0.07). We observed significant LCV results not related to brain connectivity only with respect to AgeSmk. These included brain IDPs related to cortical thickness (i.e., mean thickness of V1 in the right hemisphere generated by parcellation of the white surface using BA_exvivo parcellation, IDP 1111; gcp=0.67, P=3.33×10^−9^, rho=-0.24±0.07), regional brain volumes (i.e., volume of rostral anterior cingulate in the right hemisphere generated by parcellation of the white surface using DKT parcellation, IDP 492; gcp=0.6, P=1.45×10^−7^, rho=0.18±0.06), and cortical areas (e.g., area of orbital-inferior frontal gyrus in the right hemisphere generated by parcellation of the white surface using a2009s parcellation, IDP 958; gcp=-0.67, P=7.07×10^−10^, rho=0.22±0.08). Among LCV effects surviving FDR-significance, we observed consistent effects in the MR analyses with respect to rfMRI connectivity ICA100 edge 772 (IDP 3200, Figure 3) on DrnWk (LCV gcp=0.38, p=8.9×10^−4^, rho=-0.18±0.07; IVW beta=-0.04, 95%CI=-0.07 – −0.01). Direction consistency was observed for other 12 of the LCV significant results (Supplemental Table 5).

**Fig. 2:**
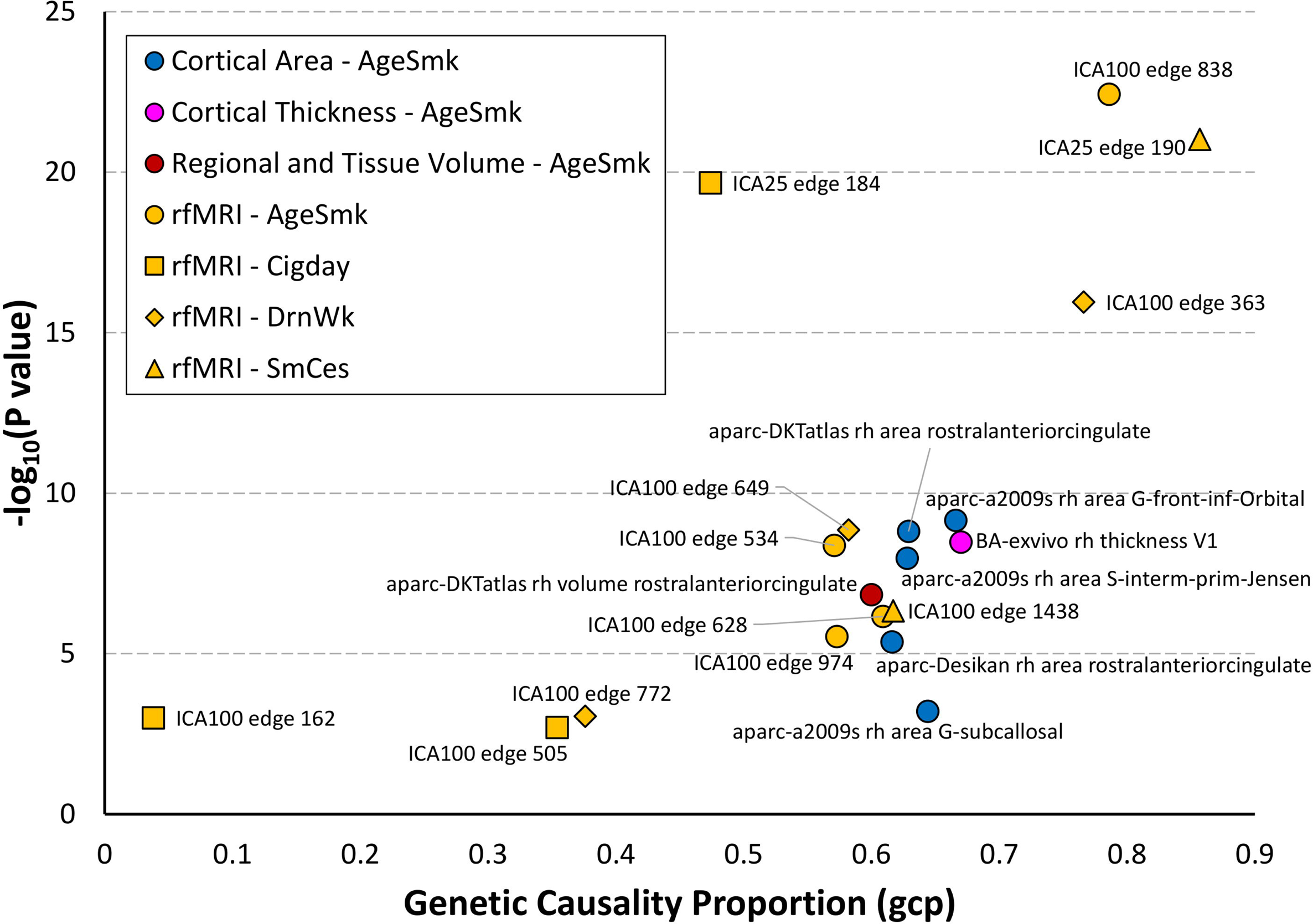
Genetic causal proportion (false discovery rate, FDR q<0.05) linking brain imaging-derived phenotypes (IDP) to alcohol drinking and tobacco smoking behaviors. Abbreviations: aparc-a2009s rh area G-front-inf-Orbital (IDP 0958); aparc-DKTatlas rh area rostralanteriorcingulate (IDP 0864); aparc-a2009s rh area S-interm-prim-Jensen (IDP 1000); aparc-Desikan rh area rostralanteriorcingulate (IDP 0707); aparc-a2009s rh area G-subcallosal (IDP 0977); BA-exvivo rh thickness V1 (IDP 1111); aparc-DKTatlas rh volume rostralanteriorcingulate (IDP 0492); ICA100 edge 838 (IDP 3266); ICA25 edge 190 (IDP 2408); ICA25 edge 184 (IDP 2402); ICA100 edge 363 (IDP 2791); ICA100 edge 649 (IDP 3077); ICA100 edge 534 (IDP 2962); ICA100 edge 1438 (IDP 3866); ICA100 edge 628 (IDP 3056); ICA100 edge 974 (IDP 3402); ICA100 edge 772 (IDP 3200); ICA100 edge 162 (IDP 2590); ICA100 edge 505 (IDP 2933). The description of brain IDPs is available in Supplemental Table 1. Full results are available in Supplemental Table 4.

**Fig. 3:**
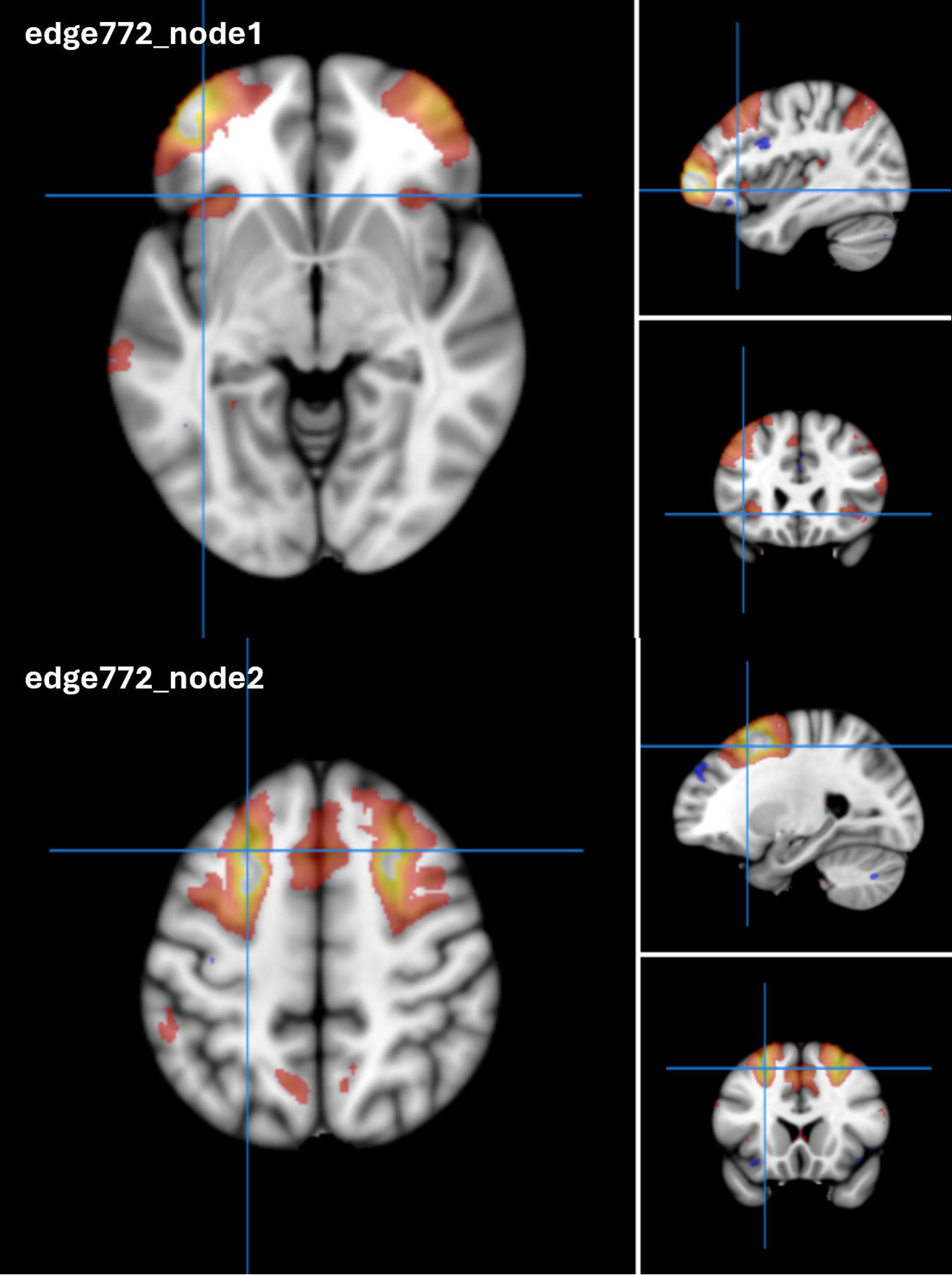
Brain imaging-derived phenotype (IDP) 3200 reflecting edge 772 of dimensionality 100 separated by spatial Independent Component Analysis (ICA) in resting-state functional magnetic resonance imaging.

## DISCUSSION

The present study uncovered new information regarding the contribution of pleiotropic mechanisms to the complex interplay of alcohol drinking and tobacco smoking with brain structure and function. Building on previous studies that reported genetically informed relationships of drinking and smoking behaviors with brain cortical morphology and grey matter volume [6,8,9], our brain-wide analyses identified genetic overlaps linking alcohol drinking and tobacco smoking with previously unexplored brain-related phenotypes, including white matter hyperintensities, specific brain substructures, and brain connectivity.

The global genetic correlation analysis identified three smoking-related results surviving multiple testing correction. Specifically, SmCes phenotype (current versus former smoker) was genetically correlated with increased total volume of white matter hyperintensities (T2 FLAIR BIANCA WMH volume, IDP 1437). Tobacco smoking has been previously linked to the progression of white matter hyperintensity in a dose-response relationship [25]. However, although this previous study did not observe an association between years since quitting tobacco smoking [25], our finding highlights a possible genetic relationship between SmCes and white matter hyperintensities. This may be due to the fact that former smokers are more likely to have quit because of health conditions and heavier tobacco use in the past [26]. Additionally, SmCes is known to be associated with long-term weight gain [27], which can lead to white matter hyperintensities via inflammation [28]. We also observed a positive genetic correlation between CigDay and the morphology of the mean second level of the right superior longitudinal fasciculus (dMRI TBSS L2 Superior longitudinal fasciculus R, IDP 1736). This brain region is part of a brain network involved in spatial awareness and proprioception [29]. Interestingly, the right superior longitudinal fasciculus has been associated with olfactory performance [30,31]. In this context, the relationship of this brain region with CigDay may be related to chemosensory processing. While there is a well-established relationship between tobacco smoking and olfactory dysfunction [32], our result suggests possible shared genetic mechanisms predisposing individuals with certain chemosensory abilities to tobacco smoking quantity. There was also a negative genetic correlation between AgeSmk and the mean thickness of pole-occipital (parc-a2009s lh thickness Pole-occipital, IDP 1219). This region is involved in visual attention [33] and appears to play a role in the reactivity toward smoking cues and tobacco craving [34,35]. In young adults, a reduced mean thickness of the left occipital pole has been associated with exposure to domestic violence [36], which is also a risk factor for tobacco smoking [37]. With respect to the relationship between AgeSmk and IDP 1219, we also observed a significant local genetic correlation in hg38 chr2:35,895,678-36,640,246 region. In this region, the GWAS catalog [23] reports a number of genome-wide significant associations, including several related to smoking initiation, chronotype, educational attainment, cognitive performance, and cortical thickness. The majority of the associations reported were related to variants mapping to *CRIM1* and *FEZ2*. *CRIM1* gene has been linked to regulatory mechanisms over axon projection targeting [38,39]. *FEZ2* is a member of a hub protein family involved in neuronal development, neurological disorders, viral infection, and autophagy [40]. While these genes were not previously linked to substance use behaviors, their functions neurodevelopment suggests mechanisms pointing to the effect of brain development on tobacco-smoking behaviors later in life.

To understand potential cause-effect relationships linking alcohol drinking and tobacco smoking to brain structure and function, we conducted a genetically informed causal inference analysis, observing convergent results between the LCV and MR methods that support an inverse effect of rfMRI connectivity ICA100 edge 772 (IDP 3200) on DrnWk phenotype. Because LCV and MR approaches are based on different assumptions, findings supported by both can be considered highly reliable. IDP 3200 reflects the activity of prefrontal and premotor cortex in the left hemisphere and that of superior frontal sulci, superior and inferior precentral sulci, and cingulate sulci (Figure 3). The left premotor cortex is involved in visual attention and in the integration of visual data at a semantic level [41,42]. Precentral and cingulate regions are associated with the modulation of reward-seeking motivation [43] and are also involved in the processing of context-related social cues [44–46]. Conversely, the right orbitofrontal region is associated with social comprehension [47]. In this context, the effect of IDP 3200 on DrnWk could reflect the impact of social cognition and decision-making process on the propensity towards alcohol consumption.

While the other LCV results did not show statistical significance in the MR analysis, we observed directional consistency for 12 of them (Supplemental Table 5). Among them, we observed that CigDay was inversely affected by fluctuations in rfMRI connectivity in edge 184 for dimensionality 25 (IDP 2402) and edge 505 for dimensionality 100 (IDP 2933). IDP 2402 reflects increased activity across left hemisphere in dorsolateral prefrontal cortex and frontal gyri, decreased activity around right orbitofrontal areas, along with increased activation of parietal lobe (Supplemental Figure 2). Being part of the superior longitudinal fasciculus, these brain areas are associated with spatial awareness and proprioception [29] and olfactory performance [30,31]. While olfactory perception is especially mediated by the orbitofrontal cortex [48], brain lesions localized within the right orbitofrontal cortex have been recorded to specifically hinder the formation of conscious olfactory percepts [49]. Interestingly, stimulating the left dorsolateral prefrontal cortex and inhibiting the right orbitofrontal cortex resulted in a risk-averse response in human subjects enrolled in a transcranial direct current stimulation study [50]. IDP 2933 reflects the association between lower activation levels in the ventral posterolateral nucleus to higher ones in the right hemisphere along the posterior cingulate cortex, dorsolateral prefrontal cortex, and frontal gyri (Supplemental Figure 3). Intriguingly, opposite fluctuation patterns were associated with increased pain in patients experiencing migraine attacks, possibly due to a strengthened network of nociceptive information processing [51]. Nicotine may have anti-nociceptive effects [52] and tobacco smoking has been linked to coping mechanisms related to migraine and chronic head pain [53]. Additionally, the interplay between nociception and olfaction has been proposed at both molecular and functional levels [54,55]. In this context, our results reinforce the hypothesis of shared genetic mechanisms predisposing individuals with certain chemosensory abilities to tobacco smoking behaviors.

In conclusion, our brain-wide analyses highlighted that different pleiotropic mechanisms likely contribute to the relationship of brain structure and function with alcohol drinking and tobacco smoking, opening new directions in understanding the processes underlying these complex behaviors. However, we also acknowledge two main limitations. While we leveraged large-scale genome-wide datasets, these were generated including only participants of European descent, because of the lack of large genetic and imaging studies in other human populations. Accordingly, our findings may not be generalizable to other population groups. Another important limitation is related to genetically informed analysis. While we used multiple methods relying on different assumptions, our results may still be affected by unaccounted confounders. Thus, our findings will need to be confirmed by evidence generated by complementary study designs (e.g., prospective studies). Finally, while no genetically inferred effect of alcohol drinking and tobacco smoking was observed, future studies with larger datasets may be able to characterize this effect direction.

## Data Availability

All data produced in the present work are contained in the manuscript

## ACKNOWLEDGEMENTS

The authors thank the participants and the investigators involved in the GWAS & Sequencing Consortium of Alcohol and Nicotine use and the UK Biobank.

## AUTHOR CONTRIBUTIONS

GD and RP conceived and designed the study. GD performed the analyses. JH and BCM provided analytic support. All authors contributed to the interpretation of the results. GD and RP drafted the manuscript. All authors edited or approved the final manuscript and agree to be accountable for its contents.

---

**Supplemental Fig. 1**: IDP 3266 reflecting edge 838 of dimensionality 100 separated by spatial ICA in resting-state functional magnetic resonance imaging.

**Supplemental Fig. 2**: IDP 2402 reflecting edge 184 of dimensionality 25 separated by spatial ICA in resting-state functional magnetic resonance imaging.

**Supplemental Fig. 3**: IDP 2933 reflecting edge 505 of dimensionality 100 separated by spatial ICA in resting-state functional magnetic resonance imaging.

## REFERENCES

1 Collaborators GBDA. Alcohol use and burden for 195 countries and territories, 1990-2016: a systematic analysis for the Global Burden of Disease Study 2016. Lancet. 2018;392(10152):1015–35.

2 He H, Pan Z, Wu J, Hu C, Bai L, Lyu J. Health Effects of Tobacco at the Global, Regional, and National Levels: Results From the 2019 Global Burden of Disease Study. Nicotine Tob Res. 2022;24(6):864–70.

3 Abburi C, Wolfman SL, Metz RA, Kamber R, McGehee DS, McDaid J. Tolerance to Ethanol or Nicotine Results in Increased Ethanol Self-Administration and Long-Term Depression in the Dorsolateral Striatum. eNeuro. 2016;3(4).

4 Morel C, Montgomery S, Han MH. Nicotine and alcohol: the role of midbrain dopaminergic neurons in drug reinforcement. Eur J Neurosci. 2019;50(3):2180–200.

5 Weera MM, Agim ZS, Cannon JR, Chester JA. Genetic correlations between nicotine reinforcement-related behaviors and propensity toward high or low alcohol preference in two replicate mouse lines. Genes Brain Behav. 2019;18(3):e12515.

6 Rabinowitz JA, Campos AI, Ong JS, Garcia-Marin LM, Alcauter S, Mitchell BL, et al. Shared Genetic Etiology between Cortical Brain Morphology and Tobacco, Alcohol, and Cannabis Use. Cereb Cortex. 2022;32(4):796–807.

7 Davey Smith G, Hemani G. Mendelian randomization: genetic anchors for causal inference in epidemiological studies. Hum Mol Genet. 2014;23(R1):R89–98.

8 Lin W, Zhu L, Lu Y. Association of smoking with brain gray and white matter volume: a Mendelian randomization study. Neurol Sci. 2023;44(11):4049–55.

9 Mavromatis LA, Rosoff DB, Cupertino RB, Garavan H, Mackey S, Lohoff FW. Association Between Brain Structure and Alcohol Use Behaviors in Adults: A Mendelian Randomization and Multiomics Study. JAMA Psychiatry. 2022;79(9):869–78.

10 Bao J, Wen J, Wen Z, Yang S, Cui Y, Yang Z, et al. Brain-wide genome-wide colocalization study for integrating genetics, transcriptomics and brain morphometry in Alzheimer’s disease. Neuroimage. 2023;280:120346.

11 He J, Cabrera-Mendoza B, De Angelis F, Pathak GA, Koller D, Curhan SG, et al. Sex differences in the pleiotropy of hearing difficulty with imaging-derived phenotypes: a brain-wide investigation. Brain. 2024.

12 Koller D, Friligkou E, Stiltner B, Pathak GA, Lokhammer S, Levey DF, et al. Pleiotropy and genetically inferred causality linking multisite chronic pain to substance use disorders. Mol Psychiatry. 2024.

13 Zanoaga MD, Friligkou E, He J, Pathak GA, Koller D, Cabrera-Mendoza B, et al. Brainwide Mendelian Randomization Study of Anxiety Disorders and Symptoms. Biol Psychiatry. 2023.

14 Saunders GRB, Wang X, Chen F, Jang SK, Liu M, Wang C, et al. Genetic diversity fuels gene discovery for tobacco and alcohol use. Nature. 2022;612(7941):720–24.

15 Smith SM, Douaud G, Chen W, Hanayik T, Alfaro-Almagro F, Sharp K, Elliott LT. An expanded set of genome-wide association studies of brain imaging phenotypes in UK Biobank. Nat Neurosci. 2021;24(5):737–45.

16 Werme J, van der Sluis S, Posthuma D, de Leeuw CA. An integrated framework for local genetic correlation analysis. Nat Genet. 2022;54(3):274–82.

17 O’Connor LJ, Price AL. Distinguishing genetic correlation from causation across 52 diseases and complex traits. Nat Genet. 2018;50(12):1728–34.

18 Bycroft C, Freeman C, Petkova D, Band G, Elliott LT, Sharp K, et al. The UK Biobank resource with deep phenotyping and genomic data. Nature. 2018;562(7726):203–09.

19 Alfaro-Almagro F, Jenkinson M, Bangerter NK, Andersson JLR, Griffanti L, Douaud G, et al. Image processing and Quality Control for the first 10,000 brain imaging datasets from UK Biobank. Neuroimage. 2018;166:400–24.

20 Bulik-Sullivan B, Finucane HK, Anttila V, Gusev A, Day FR, Loh PR, et al. An atlas of genetic correlations across human diseases and traits. Nat Genet. 2015;47(11):1236–41.

21 International HapMap C. The International HapMap Project. Nature. 2003;426(6968):789–96.

22 Genomes Project C, Auton A, Brooks LD, Durbin RM, Garrison EP, Kang HM, et al. A global reference for human genetic variation. Nature. 2015;526(7571):68–74.

23 Sollis E, Mosaku A, Abid A, Buniello A, Cerezo M, Gil L, et al. The NHGRI-EBI GWAS Catalog: knowledgebase and deposition resource. Nucleic Acids Res. 2023;51(D1):D977–D85.

24 Hemani G, Zheng J, Elsworth B, Wade KH, Haberland V, Baird D, et al. The MR-Base platform supports systematic causal inference across the human phenome. Elife. 2018;7.

25 Power MC, Deal JA, Sharrett AR, Jack CR, Jr., Knopman D, Mosley TH, Gottesman RF. Smoking and white matter hyperintensity progression: the ARIC-MRI Study. Neurology. 2015;84(8):841–8.

26 Gallus S, Muttarak R, Franchi M, Pacifici R, Colombo P, Boffetta P, et al. Why do smokers quit? Eur J Cancer Prev. 2013;22(1):96–101.

27 Veldheer S, Yingst J, Zhu J, Foulds J. Ten-year weight gain in smokers who quit, smokers who continued smoking and never smokers in the United States, NHANES 2003-2012. Int J Obes (Lond). 2015;39(12):1727–32.

28 Lampe L, Zhang R, Beyer F, Huhn S, Kharabian Masouleh S, Preusser S, et al. Visceral obesity relates to deep white matter hyperintensities via inflammation. Ann Neurol. 2019;85(2):194–203.

29 Naito E, Morita T, Amemiya K. Body representations in the human brain revealed by kinesthetic illusions and their essential contributions to motor control and corporeal awareness. Neurosci Res. 2016;104:16–30.

30 Carreiras M, Quinones I, Chen HA, Vazquez-Araujo L, Small D, Frost R. Sniffing out meaning: Chemosensory and semantic neural network changes in sommeliers. Hum Brain Mapp. 2024;45(2):e26564.

31 Segura B, Baggio HC, Solana E, Palacios EM, Vendrell P, Bargallo N, Junque C. Neuroanatomical correlates of olfactory loss in normal aged subjects. Behav Brain Res. 2013;246:148–53.

32 Ajmani GS, Suh HH, Wroblewski KE, Pinto JM. Smoking and olfactory dysfunction: A systematic literature review and meta-analysis. Laryngoscope. 2017;127(8):1753–61.

33 Mevorach C, Hodsoll J, Allen H, Shalev L, Humphreys G. Ignoring the elephant in the room: a neural circuit to downregulate salience. J Neurosci. 2010;30(17):6072–9.

34 Mondino M, Luck D, Grot S, Januel D, Suaud-Chagny MF, Poulet E, Brunelin J. Effects of repeated transcranial direct current stimulation on smoking, craving and brain reactivity to smoking cues. Sci Rep. 2018;8(1):8724.

35 Yang Z, Zhang Y, Cheng J, Zheng R. Meta-analysis of brain gray matter changes in chronic smokers. Eur J Radiol. 2020;132:109300.

36 Tomoda A, Polcari A, Anderson CM, Teicher MH. Reduced visual cortex gray matter volume and thickness in young adults who witnessed domestic violence during childhood. PLoS One. 2012;7(12):e52528.

37 Budenz A, Klein A, Prutzman Y. The Relationship Between Trauma Exposure and Adult Tobacco Use: Analysis of the National Epidemiologic Survey on Alcohol and Related Conditions (III). Nicotine Tob Res. 2021;23(10):1716–26.

38 Sahni V, Itoh Y, Shnider SJ, Macklis JD. Crim1 and Kelch-like 14 exert complementary dual-directional developmental control over segmentally specific corticospinal axon projection targeting. Cell Rep. 2021;37(3):109842.

39 Sahni V, Shnider SJ, Jabaudon D, Song JHT, Itoh Y, Greig LC, Macklis JD. Corticospinal neuron subpopulation-specific developmental genes prospectively indicate mature segmentally specific axon projection targeting. Cell Rep. 2021;37(3):109843.

40 Teixeira MB, Alborghetti MR, Kobarg J. Fasciculation and elongation zeta proteins 1 and 2: From structural flexibility to functional diversity. World J Biol Chem. 2019;10(2):28–43.

41 Hertrich I, Dietrich S, Blum C, Ackermann H. The Role of the Dorsolateral Prefrontal Cortex for Speech and Language Processing. Front Hum Neurosci. 2021;15:645209.

42 Bartel G, Marko M, Rameses I, Lamm C, Riecansky I. Left Prefrontal Cortex Supports the Recognition of Meaningful Patterns in Ambiguous Stimuli. Front Neurosci. 2020;14:152.

43 Dubey I, Georgescu AL, Hommelsen M, Vogeley K, Ropar D, Hamilton AFC. Distinct neural correlates of social and object reward seeking motivation. Eur J Neurosci. 2020;52(9):4214–29.

44 Apps MA, Rushworth MF, Chang SW. The Anterior Cingulate Gyrus and Social Cognition: Tracking the Motivation of Others. Neuron. 2016;90(4):692–707.

45 Gordon EM, Chauvin RJ, Van AN, Rajesh A, Nielsen A, Newbold DJ, et al. A somato-cognitive action network alternates with effector regions in motor cortex. Nature. 2023;617(7960):351–59.

46 Lavin C, Melis C, Mikulan E, Gelormini C, Huepe D, Ibanez A. The anterior cingulate cortex: an integrative hub for human socially-driven interactions. Front Neurosci. 2013;7:64.

47 Nakamura M, Nestor PG, Shenton ME. Orbitofrontal Sulcogyral Pattern as a Transdiagnostic Trait Marker of Early Neurodevelopment in the Social Brain. Clin EEG Neurosci. 2020;51(4):275–84.

48 Sagar V, Shanahan LK, Zelano CM, Gottfried JA, Kahnt T. High-precision mapping reveals the structure of odor coding in the human brain. Nat Neurosci. 2023;26(9):1595–602.

49 Li W, Lopez L, Osher J, Howard JD, Parrish TB, Gottfried JA. Right orbitofrontal cortex mediates conscious olfactory perception. Psychol Sci. 2010;21(10):1454–63.

50 Nejati V, Salehinejad MA, Nitsche MA. Interaction of the Left Dorsolateral Prefrontal Cortex (l-DLPFC) and Right Orbitofrontal Cortex (OFC) in Hot and Cold Executive Functions: Evidence from Transcranial Direct Current Stimulation (tDCS). Neuroscience. 2018;369:109–23.

51 Lim M, Jassar H, Kim DJ, Nascimento TD, DaSilva AF. Differential alteration of fMRI signal variability in the ascending trigeminal somatosensory and pain modulatory pathways in migraine. J Headache Pain. 2021;22(1):4.

52 Carstens E, Carstens MI. Sensory Effects of Nicotine and Tobacco. Nicotine Tob Res. 2022;24(3):306–15.

53 Weinberger AH, Seng EK. The Relationship of Tobacco Use and Migraine: A Narrative Review. Curr Pain Headache Rep. 2023;27(4):39–47.

54 Mignot C, Faria V, Hummel T, Frost M, Michel CM, Gossrau G, Haehner A. Migraine with aura: less control over pain and fragrances? J Headache Pain. 2023;24(1):55.

55 Lotsch J, Hahner A, Gossrau G, Hummel C, Walter C, Ultsch A, Hummel T. Smell of pain: intersection of nociception and olfaction. Pain. 2016;157(10):2152–57.

