## Supplemental Figures for "Brain-wide pleiotropy investigation of alcohol drinking and tobacco smoking behaviors"

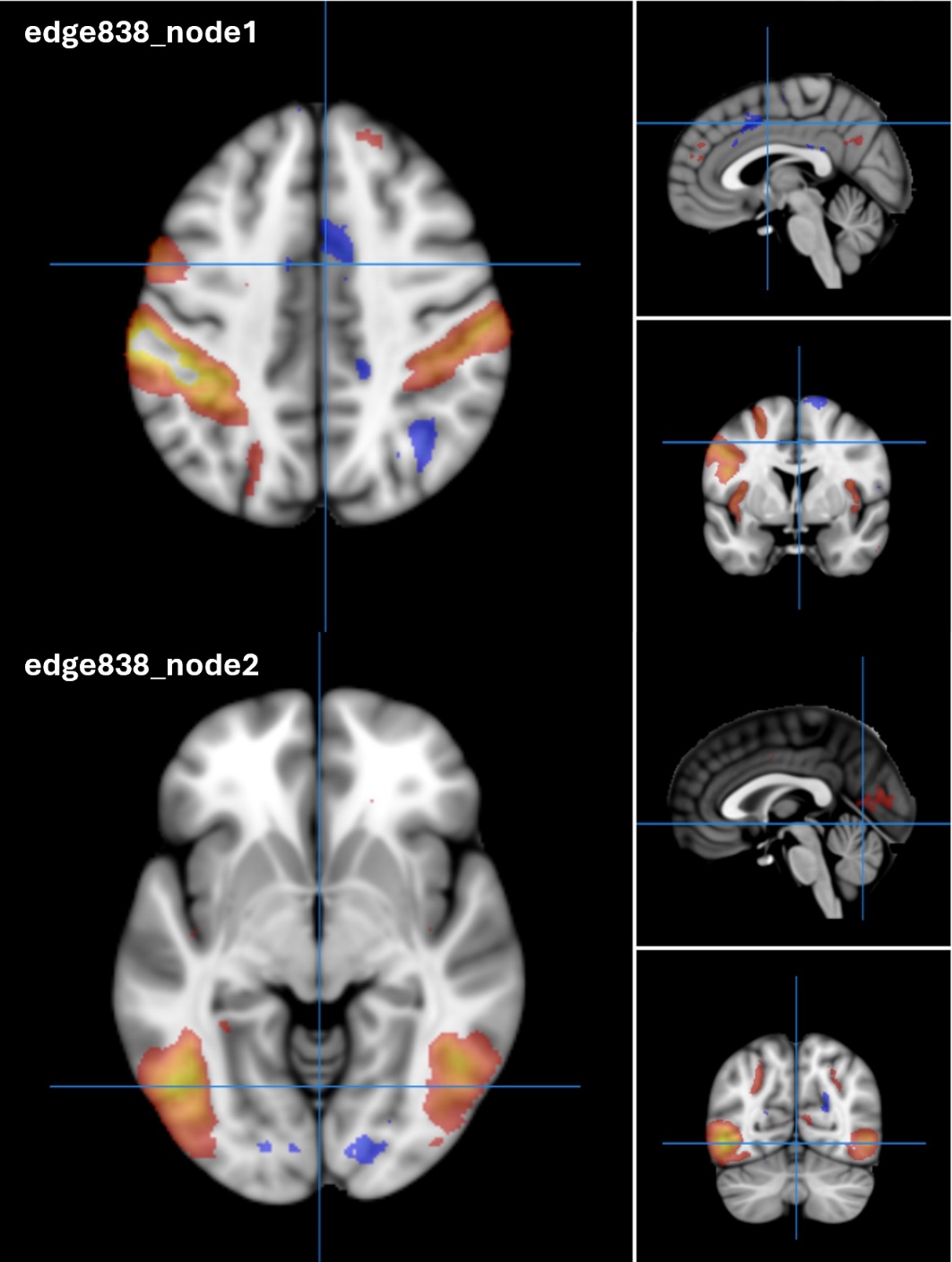

**Supplemental Fig. 1**: Brain imaging-derived phenotypes (IDP) 3266 reflecting edge 838 of dimensionality 100 separated by spatial Independent Component Analysis (ICA) in resting-state functional magnetic resonance imaging.

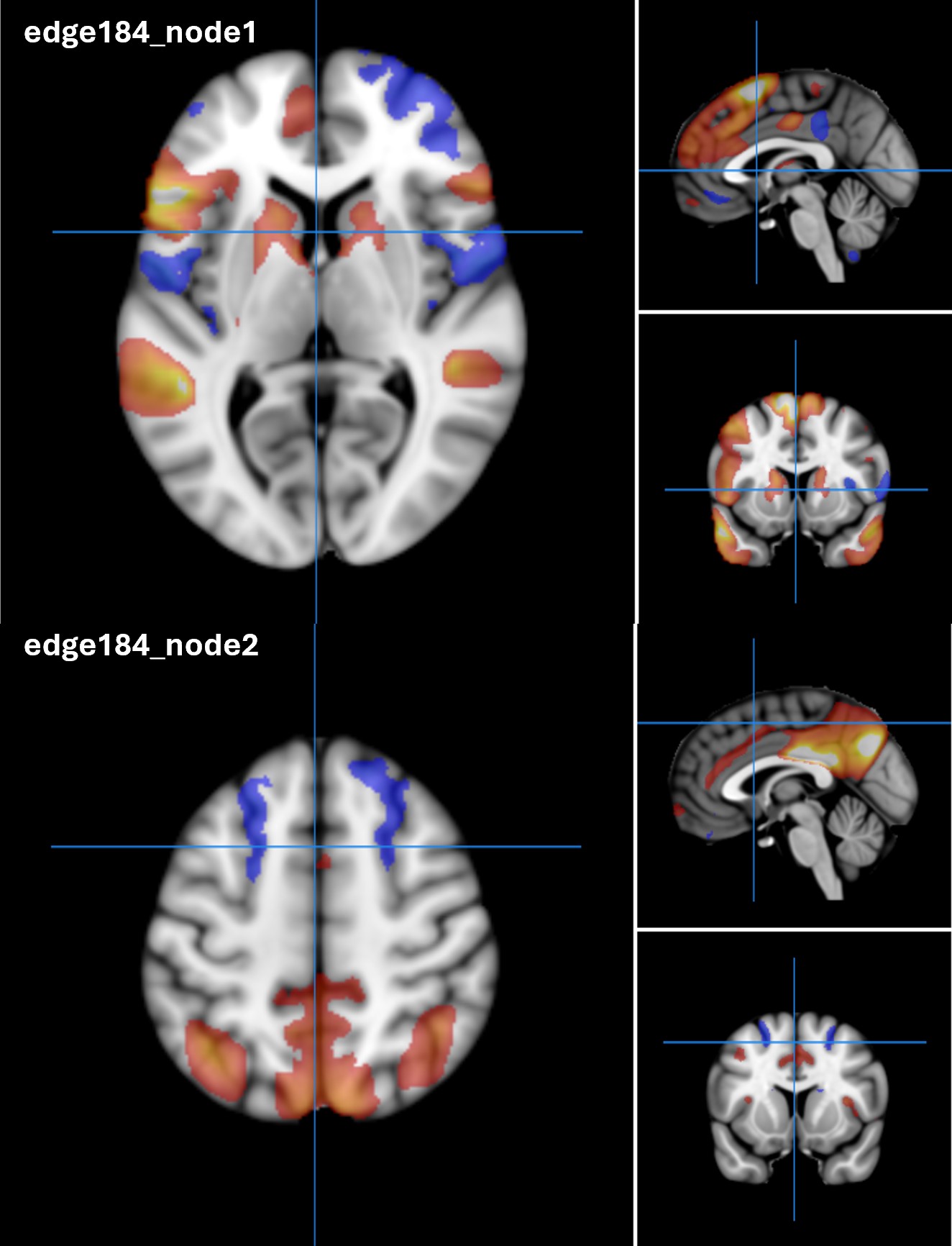

**Supplemental Fig. 2**: IDP 2402 reflecting edge 184 of dimensionality 25 separated by spatial ICA in resting-state functional magnetic resonance imaging.

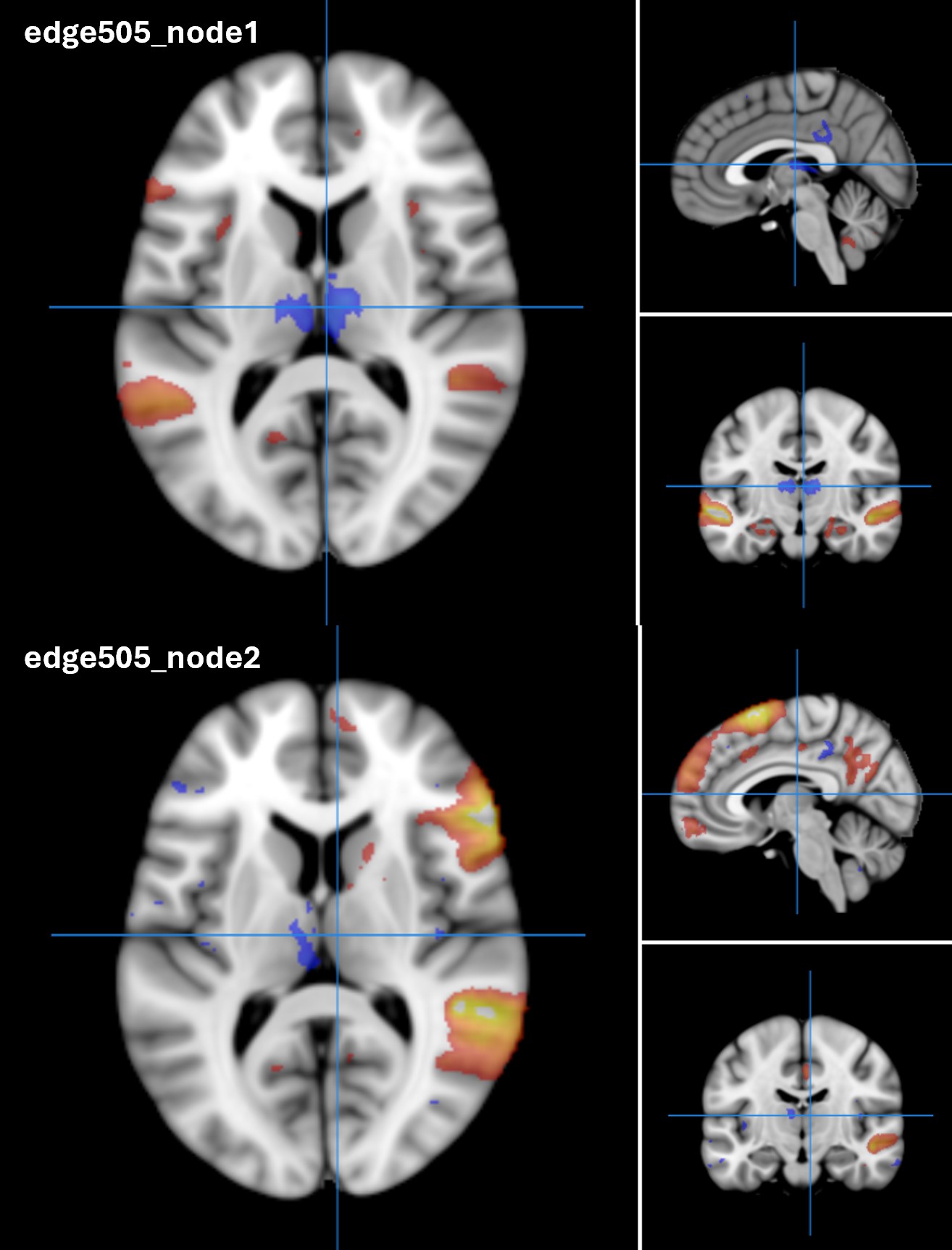

**Supplemental Fig. 3**: IDP 2933 reflecting edge 505 of dimensionality 100 separated by spatial ICA in resting-state functional magnetic resonance imaging.
